# The Risk Factors, Detection and Classification of Esophageal Cancer Using Ensemble Machine Learning Models

**DOI:** 10.64898/2026.03.09.26347944

**Authors:** Mekia Shigute Gaso, Remudin Reshid Mekuria, Selcuk Cankurt, Haji Aman Deybasso, Abdella Amano Abdo, Ghulam Husain Abbas

**Affiliations:** Computer Science Department, Ala-Too International University, Bishkek, Kyrgyzstan; Applied Mathematics and Informatics Department, Ala-Too International University, Bishkek, Kyrgyzstan; Computer Engineering Department, Vistula University, Warsaw, Poland; Department of Public Health, Adama Hospital Medical College, Adama, Ethiopia; Research and Community Service Office, Negelle Arsi General Hospital and Medical College, Negelle Arsi, Ethiopia; Faculty of Medicine, Ala-Too International University, Bishkek, Kyrgyzstan

**Keywords:** Esophageal cancer, Machine learning, Ensemble learning, Feature selection, Gradient boosting, XGBoost, Random Forest, Medical classification

## Abstract

Esophageal cancer (EC) remains one of the most lethal malignancies worldwide, with poor survival outcomes largely attributable to late-stage diagnosis and limited treatment effectiveness. Early detection and accurate risk stratification are therefore essential for improving clinical management. In this study, we investigate the predictive value of socio-demographic, dietary, behavioral, environmental, and clinical variables collected from 312 individuals (104 EC cases and 208 controls) in the Arsi Zone, Ethiopia.

An ensemble features ranking approach based on Random Forest machine learning was first applied to identify the most relevant predictive features. Subsequently, multiple ensemble machine learning models were evaluated, including Histogram-based Gradient Boosting (Model I), Extreme Gradient Boosting (Model II), AdaBoost (Model III), Random Forest (Model IV), and k-Nearest Neighbors (Model V). These models were tested under multiple experimental settings using both full and reduced feature subsets. To enhance robustness and minimize variability, a multi-seed ensemble framework was employed. Different seed values generate distinct train–test splits and slight variations in model initialization and optimization, leading to minor differences in training outcomes; aggregating results across multiple seeds mitigates this variability and provides more stable and reliable performance estimates.

The experimental results demonstrate that boosting-based ensemble models consistently outperform other classifiers across all evaluation metrics. Model I achieved the highest overall performance, reaching an accuracy of 0.983, with precision of 0.982, recall of 0.980, and F1-score of 0.981 using the reduced feature set, while maintaining nearly identical performance with the full feature set. Model II also showed stable and strong predictive capability, achieving accuracies of 0.963 and 0.961 for the full and reduced feature sets, respectively, with balanced precision, recall, and F1-score values. These findings indicate that feature importance-based dimensionality reduction preserves essential predictive information without compromising classification performance.

Overall, the results highlight the significant predictive contribution of dietary and environmental risk factors and demonstrate that ensemble learning provides a reliable, efficient, and clinically meaningful approach for early EC detection. The proposed framework offers a promising direction for supporting diagnostic decision-making and risk stratification in resource-limited healthcare settings.

**Highlights:**

1. Machine Learning Framework for Esophageal Cancer Classification A robust ensemble machine learning framework was developed to classify esophageal cancer using socio-demographic, dietary, behavioral, environmental, and clinical risk factors, enabling accurate and reliable disease prediction.
2. Multi-Seed Ensemble Strategy for Improved Model Stability A novel multi-seed ensemble classification approach was implemented to reduce model variance and improve robustness by aggregating predictions across multiple randomized training and testing splits.
3. Ensemble Feature Ranking for Optimal Feature Selection An ensemble Random Forest–based feature ranking framework was designed to identify the most predictive features, ensuring stable biomarker selection and improved model interpretability.
4. High Classification Performance with Reduced Feature Set The proposed ensemble HGBC model achieved outstanding performance with 98.3% accuracy, 98.2% precision, 98.0% recall, and 98.1% F1-score using a reduced feature subset, demonstrating efficient dimensionality reduction without performance loss.
5. Exceptional Discriminative Ability with Near-Perfect AUC The ensemble HGBC model achieved an AUC of 0.994, indicating excellent discrimination between cancer and non-cancer cases and confirming its suitability for high-precision clinical decision support.
6. Zero False-Negative Predictions and Maximum Diagnostic Sensitivity The proposed model achieved zero false negatives in evaluation, resulting in 100% statistical power and perfect sensitivity, ensuring reliable detection of esophageal cancer cases.
7. Identification of Key Dietary and Environmental Risk Factors Feature importance analysis revealed that dietary habits, hot food consumption, environmental exposures, and behavioral factors are among the most significant predictors of esophageal cancer risk.
8. Ensemble Learning Outperforms Traditional Machine Learning Models Boosting-based ensemble models, particularly HGBC and XGBoost, consistently outperformed other classifiers, demonstrating superior predictive accuracy, stability, and robustness.
9. Efficient and Interpretable AI Framework for Clinical Decision Support The proposed framework balances high predictive accuracy with interpretability, making it suitable for assisting clinicians in early diagnosis and risk stratification of esophageal cancer.
10. AI-Driven Solution for Resource-Constrained Healthcare Settings The proposed ensemble machine learning approach provides an effective and scalable diagnostic support tool, particularly valuable for healthcare systems with limited resources and access to specialized medical expertise.

## 1. Introduction

Esophageal cancer (EC) is a highly lethal malignancy characterized by abnormal cell growth in the esophageal lining. It remains a major global health concern, ranking as the seventh most common cancer worldwide and the sixth leading cause of cancer-related deaths [1]. Approximately 570,000 new cases are reported annually, with particularly high prevalence in regions such as East Asia and Eastern and Southern Africa, where esophageal squamous cell carcinoma (ESCC) is the predominant subtype [2]. In contrast, esophageal adenocarcinoma (EAC), the other major histological type, is more common in Western countries, including the United States and Europe. Both subtypes exhibit significant regional variation due to differences in environmental exposure, lifestyle behaviors, and genetic susceptibility [3].

The etiology of EC varies according to its histological subtype. ESCC is strongly associated with lifestyle-related risk factors, including tobacco use, alcohol consumption, nutritional deficiencies, and frequent intake of hot beverages. Conversely, EAC is primarily linked to obesity, gastroesophageal reflux disease (GERD), and Barrett’s esophagus. In addition, genetic predisposition and environmental exposures contribute significantly to the development of both ESCC and EAC. Recent studies highlight that genetic mutations, poor dietary habits, and environmental risk factors play a critical role in the high incidence of ESCC in endemic regions [4].

Despite advances in medical treatment, EC remains a challenging malignancy with poor prognosis, largely due to late-stage diagnosis. The five-year survival rate remains below 15%, even with current treatment approaches such as surgery, chemotherapy, and radiotherapy [5]. However, recent developments in targeted therapy and immunotherapy offer promising opportunities to improve patient outcomes. Immunotherapy, particularly immune checkpoint inhibitors, has shown encouraging preliminary results in EC treatment. In addition, adoptive cell therapy and cancer vaccine research have demonstrated potential in clinical trials [6]. Although immunotherapy has emerged as an important treatment option for advanced EC, several challenges remain, including identifying patients who will benefit most, managing immune-related adverse effects, and optimizing combination strategies with conventional treatments [6].

Given the complexity and multifactorial nature of EC risk factors, there is an urgent need for robust and scalable approaches to improve early detection and risk stratification. Machine learning (ML) techniques provide powerful tools for analyzing large and complex datasets, identifying hidden patterns, and improving diagnostic accuracy. By integrating clinical, demographic, behavioral, and environmental data, ML models can assist in identifying high-risk individuals and supporting early diagnosis. This approach has the potential to enhance screening programs, improve clinical decision-making, and enable personalized treatment planning. Therefore, this study aims to explore the application of ensemble machine learning techniques to identify key risk factors and develop accurate predictive models for esophageal cancer detection and classification.

The remainder of this paper is organized as follows. Section 2 presents a review of related work. Section 3 describes the research methodology. Section 4 introduces the ensemble-based machine learning framework, including feature ranking and multi-seed classification strategies. Section 5 presents experimental results and discussion. Finally, Section 6 concludes the paper and provides recommendations for future research.

## 2. Literature Review

Esophageal cancer (EC) is a multifactorial disease influenced by a wide variety of environmental, lifestyle, and genetic determinants. Although numerous studies have examined its etiology, the relative contribution and interaction of these factors remain only partially understood. Early diagnosis continues to play a crucial role in reducing disease burden, and white-light endoscopy remains the primary clinical tool for screening and early lesion detection [7].

A large body of epidemiological work has investigated the risk profile of esophageal squamous cell carcinoma (ESCC), though consensus on its dominant causative pathways is still evolving. Foundational studies have examined major behavioral and environmental exposures and assessed their magnitude from a public-health standpoint [8]. In regions forming the traditional “esophageal cancer belt,” including parts of central and northern China, research consistently points to inadequate fruit and vegetable consumption, tobacco and alcohol use, nutritional deficiencies, and habitual intake of scalding beverages as central contributors to the high regional incidence of ESCC [9,10]. Other conditions, such as chronic gastrointestinal disorders, head and neck cancers, human papillomavirus (HPV) infection, inherited syndromes like tylosis, low socioeconomic status, and suboptimal oral hygiene, also appear to elevate risk, though their significance varies geographically [11,12].

Broader epidemiological analyses reflect similar patterns. Lifestyle-related exposures such as smoking, consumption of very hot tea, red-meat–dominant diets, and poor oral health practices have repeatedly been associated with increased ESCC risk, while low intake of fresh produce and micronutrient-dense foods further exacerbates susceptibility [13]. Esophageal adenocarcinoma (EAC), on the other hand, follows a different etiological trajectory: Barrett’s esophagus serves as its primary precursor, with dysplastic changes representing the strongest predictor of malignant progression. Recent trends in mortality, especially for EAC, underscore the need for deeper investigation into its pathogenesis and improved targeted prevention strategies [7].

Environmental exposures, dietary habits, and socioeconomic inequalities all shape EC risk across global populations [8]. High-risk geographic regions share common patterns of nutritional insufficiency, widespread tobacco use, and cultural consumption of extremely hot beverages, contributing to distinct disease burdens [9,10]. Additional susceptibility arises from underlying medical conditions, viral infections, and genetically mediated vulnerabilities, further highlighting the complexity of EC etiology [11,12]. Strengthening understanding of how these interacting factors influence disease development remains essential for improving screening and prevention strategies.

Nutritional influences constitute another important domain within EC research. Numerous studies suggest that diets rich in fruits, vegetables, antioxidants, folate, vitamin C, and fiber exert a protective effect against both ESCC and EAC [14–16]. Conversely, heavy alcohol consumption, particularly spirits and beer, and tobacco use substantially increase EC risk, with synergistic carcinogenic effects observed when these exposures co-occur [17–19]. Smoking continues to be one of the strongest predictors of ESCC, and its adverse impact persists long after cessation [20,21].

Beyond environmental and behavioral influences, gastroesophageal reflux disease (GERD) is a major determinant of EAC. Recurrent exposure of the esophageal epithelium to gastric acid can lead to metaplastic changes, ultimately promoting malignant transformation [22]. Genetic and familial studies further indicate that EC may follow an autosomal-recessive inheritance pattern in certain populations, although the interplay between genetic architecture and environmental triggers remains insufficiently characterized [23,24]. Recent advances in genomics and epigenetics have identified key molecular abnormalities, including mutations and regulatory alterations, that offer potential for earlier detection and more individualized therapeutic approaches [25,26].

Taken together, the literature characterizes EC as a disease arising from the interplay of diverse risk factors spanning behavior, diet, environment, and genetic predisposition. This growing body of evidence highlights the importance of comprehensive prevention efforts, ranging from lifestyle modification to improved screening practices, as well as continued research aimed at elucidating the biological mechanisms that drive esophageal carcinogenesis.

## 3. Research Methodology

### 3.1. Data Acquisition

Data were collected using close-ended questionnaires (checklists) that comprised demographic characteristics (age, sex, and residence), clinical, histopathological type of cancer, tumor location, degree of tumor differentiation, cancer stages, and the primary residences of the patients. The severity of dysphagia was graded as follows: grade 1: normal swallowing; grade 2: difficulty in swallowing solids; grade 3: difficulty in swallowing semisolids; grade 4: difficulty in swallowing liquids; grade 5: difficulty in swallowing own saliva. The clinical staging was performed using TNM (American Joint Committee on Cancer (AJCC) cancer staging manual); this is a staging system that is an expression of the anatomical extent of the disease based on the extent of the primary tumor (T), absence or presence of and extent of regional lymph node metastasis (N) and absence or presence of distant metastasis (M). The administrative units, geographical locations, and agro-climatic divisions were gathered from the Arsi Zone finance and economic development office, while the topologies of the study area were collected by using Geographical Information System (GIS) obtained from the agriculture departments of Arsi Zone.

The dataset comprised 312 instances, of which 208 were controls and 104 were confirmed esophageal cancer cases, and included predictors spanning socio-demographic, dietary, behavioral, environmental, and clinical domains.

### 3.2. Pre-processing of the Data

As part of the pre-processing workflow, columns corresponding to unique identifiers were removed because they do not provide any predictive value for the classification task. In addition, a comprehensive exploratory data analysis was performed, building on our earlier visualization-focused work in [27]. Missing values (NaNs), which typically arise from incomplete data entry or unavailable measurements, were eliminated to ensure consistency and reliability for downstream modeling. After completing all data cleaning steps, the final working dataset consisted of 312 instances and 52 relevant features, which are hereafter referred to as the full feature set. This cleaned and reduced dataset was then partitioned into training (80%) and validation (20%) subsets.

### 3.3. Implementation of the Machine Learning Models

Machine learning (ML) is a branch of artificial intelligence (AI) that focuses on developing algorithms capable of learning patterns from data and making predictions or decisions without explicit programming. ML techniques can be broadly categorized into supervised, unsupervised, and reinforcement learning, each serving distinct purposes based on the nature of the available data. Supervised learning uses labeled data to train models for tasks such as classification and regression, while unsupervised learning identifies hidden structures in unlabeled data. With the rapid advancements in computing power and the availability of large datasets, machine learning has been successfully applied in diverse fields such as healthcare, finance, and autonomous systems, driving innovation and automation across industries.

The following machine learning models were implemented and evaluated using the dataset:

**A. Histogram-based Gradient Boosting Classifier (HGBC)** is an efficient implementation of gradient boosting that improves computational performance by discretizing continuous features into histograms before training [28]. Unlike traditional gradient boosting, which evaluates each data point individually, HGBC groups feature values into discrete bins, significantly reducing memory consumption and training time. The model sequentially builds an ensemble of weak learners, typically decision trees, where each new tree is trained to correct the residual errors of the previous ones. At each iteration, the model updates its predictions using the additive model:

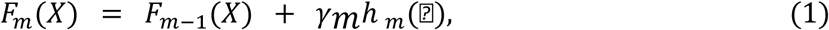

where (*F*_*m*_(*X*)) is the ensemble model at iteration (*m*), (*h* _*m*_(⍰)) is the weak learner, and (*γ*_*m*_) is the learning rate. The algorithm minimizes a differentiable loss function, such as squared error or log loss, through gradient descent. Our HGBC model is implemented with 100 iterations, a learning rate of 0.1, and histogram-based binning for computational efficiency.

**B. Adaptive Boosting (AdaBoost)** is an ensemble algorithm that combines multiple weak classifiers into a strong classifier by iteratively reweighting misclassified samples [29]. Unlike bagging-based methods, which train classifiers independently on bootstrapped samples, AdaBoost assigns higher weights to difficult-to-classify instances, forcing subsequent weak learners to focus on these challenging points. The final prediction is obtained through a weighted combination of all weak learners:

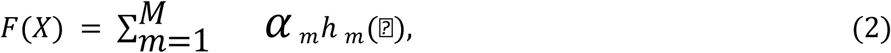

where (*h* _*m*_(⍰)) represents the weak classifiers, (*⍺* _*m*_) is the weight assigned to classifier (*m*), and (*M*) is the total number of boosting rounds. AdaBoost minimizes an exponential loss function, which increases the emphasis on misclassified data points. Our implementation uses 50 estimators and a learning rate of 1.0 to balance model flexibility and generalization.

**C. k-Nearest Neighbors (KNN)** is a non-parametric, instance-based learning algorithm that classifies new samples based on their similarity to existing data. For a given observation, the algorithm identifies the (K) closest training points using a chosen distance metric, commonly the Euclidean distance, and assigns a class label by majority vote [30]. Since KNN does not learn an explicit model, computational cost is incurred during prediction, where distances between the new instance and all training samples must be computed. The classification decision is given by:

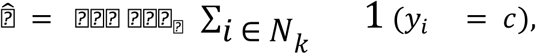

where ( *N*_*k*_) denotes the set of (*k*) nearest neighbors and 1(⋅) is an indicator function [31]. Our KNN model uses (K = 100), providing smoother decision boundaries and reducing sensitivity to noise.

**D. Extreme Gradient Boosting (XGBoost)** is a highly optimized gradient boosting framework designed for efficiency, scalability, and regularized learning [32]. XGBoost employs second-order gradient information, shrinkage, and column subsampling to improve model accuracy while preventing overfitting. It also introduces a regularized objective function:

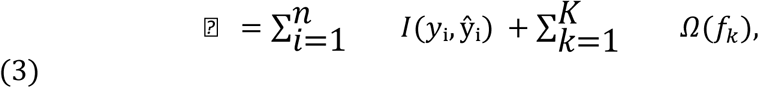

where *L(·)* is a differentiable loss function and Ω(*f*_*k*_) penalizes the complexity of individual trees. This regularization distinguishes XGBoost from classical gradient boosting methods and improves generalization performance. Additional optimizations, such as sparsity-aware learning and parallel tree construction, enable fast model training even on large datasets. In this study, XGBoost achieved consistently high performance on both full and reduced feature sets.

**E. Random Forest (RF)** is an ensemble learning method that constructs a collection of decision trees, each trained on a bootstrap sample of the data and a random subset of features [33]. By combining predictions from many decorrelated trees, Random Forest reduces variance and mitigates overfitting. The prediction for classification tasks is obtained through majority voting across all trees. Formally, for “T” trees, the classifier output is:

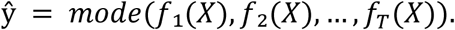

The random feature selection at each split ensures that the trees are diverse, which strengthens the ensemble’s generalization ability. Random Forests also provide feature importance estimates based on impurity reduction or permutation scores. In our experiments, the RF model offered competitive accuracy, particularly in the ensemble configuration.

## 4. Ensemble Framework

This section outlines the ensemble-based strategies integrated into our machine learning pipeline, including the ensemble feature-ranking procedure and the multi-seed ensemble classification framework, both of which are designed to enhance model stability and robustness. These components are discussed briefly in the following subsections.

### 4.1 Ensemble Feature Ranking Framework

We designed an ensemble feature-ranking framework, illustrated in Fig. 1. To enhance the stability of biomarker identification, we extended the multi-seed strategy to a Random Forest (RF)-based feature-ranking model. Each RF model, configured using different seed values and trained on a corresponding seed-specific train–test split, produced an independent ranking of feature importance. Single-run feature importance scores are known to fluctuate in biomedical settings due to correlations between variables and limited sample sizes [34]. Therefore, analyzing importance values across multiple seeded models and training data splits provides a more reliable assessment of each feature’s contribution and reduces the risk of identifying spurious biomarkers.

**Figure 1.**
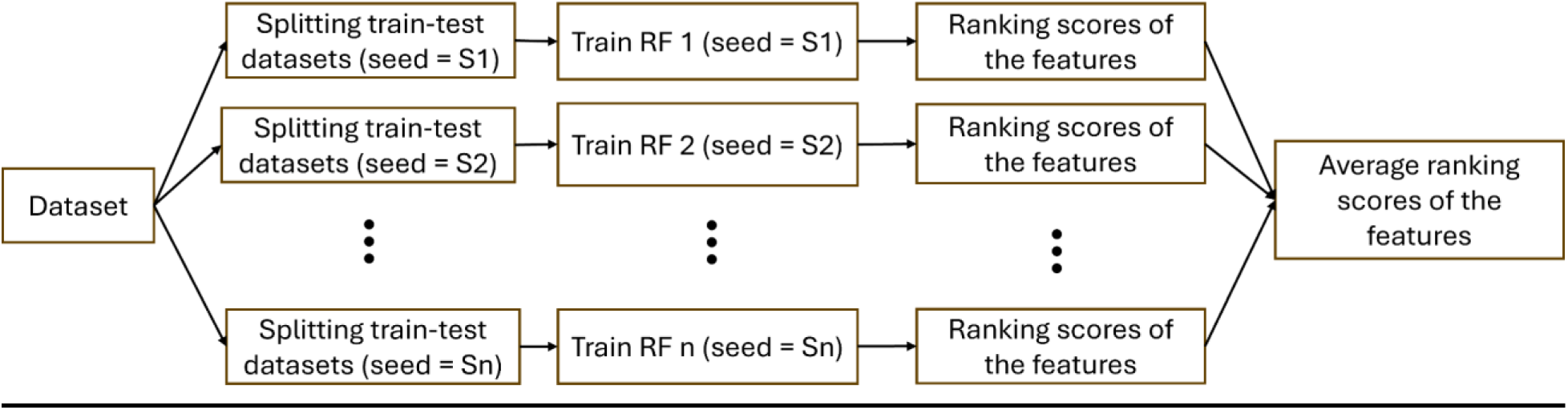
A chart of an ensemble feature ranking model framework.

The final feature importance profile was obtained by averaging importance scores across all seed-specific RF models, resulting in a more reliable and robust ranking [35, 36]. Ensemble-based feature aggregation has been recommended in prior studies to enhance the stability of feature selection and improve interpretability in biomedical research [33, 37]. For esophageal cancer applications, such stability is crucial for highlighting features, whether clinical, molecular, or imaging-based, that consistently contribute to classification performance. This helps ensure that the identified biomarkers have stronger biological and translational relevance.

### 4.2 Ensemble Classification Framework

In Fig. 2, we designed a framework for the machine learning ensemble classifier. To ensure robust model performance, we employed a multi-seed homogeneous ensemble classification strategy, as shown in Fig. 2. The dataset was repeatedly split using different random seeds (S₁, S₂, …, Sₙ), and the same base learners were configured using different random seeds (S₁, S₂, …, Sₙ), which were then trained on each corresponding seed-specific partition. This approach minimizes the dependence of the model on a single train–test split and reduces variance, an important consideration given the heterogeneity and typically limited sample sizes found in esophageal cancer datasets [38, 39]. By training multiple models across different data segments, the ensemble captures a broader representation of the underlying data distribution, aligning with established ensemble learning theory [40].

**Figure 2.**
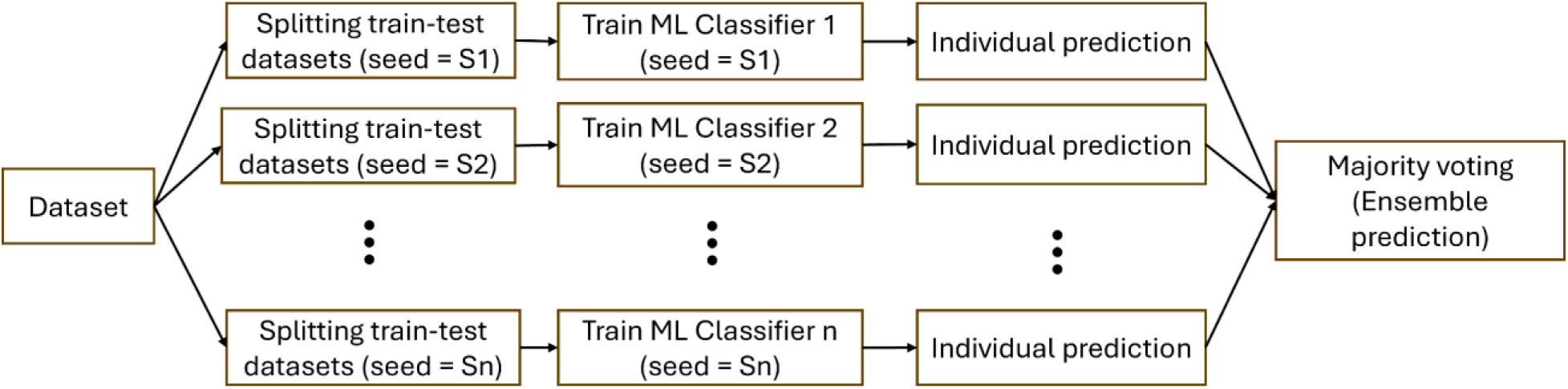
A chart of a machine learning ensemble classifier framework.

Following training, each classifier provided individual predictions, which were aggregated using majority voting to obtain the final ensemble decision. Majority voting is a simple yet effective method for reducing the influence of outlying predictions and improving classification stability [41]. In medically oriented classification problems, such as early detection or subtype identification in esophageal cancer, ensemble methods have demonstrated improved reliability and robustness compared with single-model approaches [42, 43]. Thus, the multi-seed ensemble contributes to a more stable and clinically relevant diagnostic pipeline.

HGBC introduces randomness through histogram binning and subsampling strategies, which diversify the weak learners and stabilize training. AdaBoost, while primarily deterministic in its weight-updating procedure, often relies on randomized base learners such as decision stumps or shallow trees to generate variation across iterations. XGBoost extends gradient boosting with additional stochastic elements, including column subsampling and randomization in tree construction, thereby improving efficiency and robustness. Random Forests explicitly embed randomness through bootstrap sampling of the training data and random feature selection at each split, ensuring high diversity among trees and yielding strong variance reduction.

In contrast, KNN is a deterministic algorithm. When provided with identical training data and hyperparameters, it consistently produces the same predictions, with no stochastic variation. For this reason, our proposed ensemble framework is only able to introduce diversity by applying random training and test splits across individual KNN models. Other models, however, benefit from randomization both in data partitioning and in the configuration of model stages within our proposed ensemble design. In contrast, ensemble and boosting methods typically incorporate randomness to enhance generalization and reduce overfitting. Together, these stochastic mechanisms help the proposed ensemble models mitigate the effects of randomness, resulting in more reliable predictive performance.

## 5. Results and Discussion

In this section, we present and analyze the predictive performance of the ensemble models designed based on the machine learning algorithms described in Section 3.3. We first examine feature importance to identify the key socio-demographic, dietary, behavioral, and environmental variables contributing to esophageal cancer risk. We then evaluate classification performance using multiple metrics, including accuracy, precision, recall, F1-score, and ROC curves, across both full and reduced feature sets.

Two experimental setups were conducted to assess the impact of feature reduction on model effectiveness, and ensemble classifiers were compared to evaluate robustness and stability. The discussion highlights the interplay between biologically plausible risk factors and model performance, providing insight into the practical utility of these predictive frameworks in clinical and resource-limited settings.

Fig. 3 shows the feature-importance profile from the ensemble feature ranking model. It highlights a cluster of dietary and environmental variables as the most influential predictors, a pattern that is consistent with established epidemiological evidence on esophageal cancer risk. Highly ranked features such as “Q302Fsweets,” “Q297Folisandfats,” “Q312FCondiments,” and radiation exposure align with research linking high intake of sugary foods, unhealthy fats, and preserved or condiment-rich diets with elevated esophageal cancer risk.

**Figure 3.**
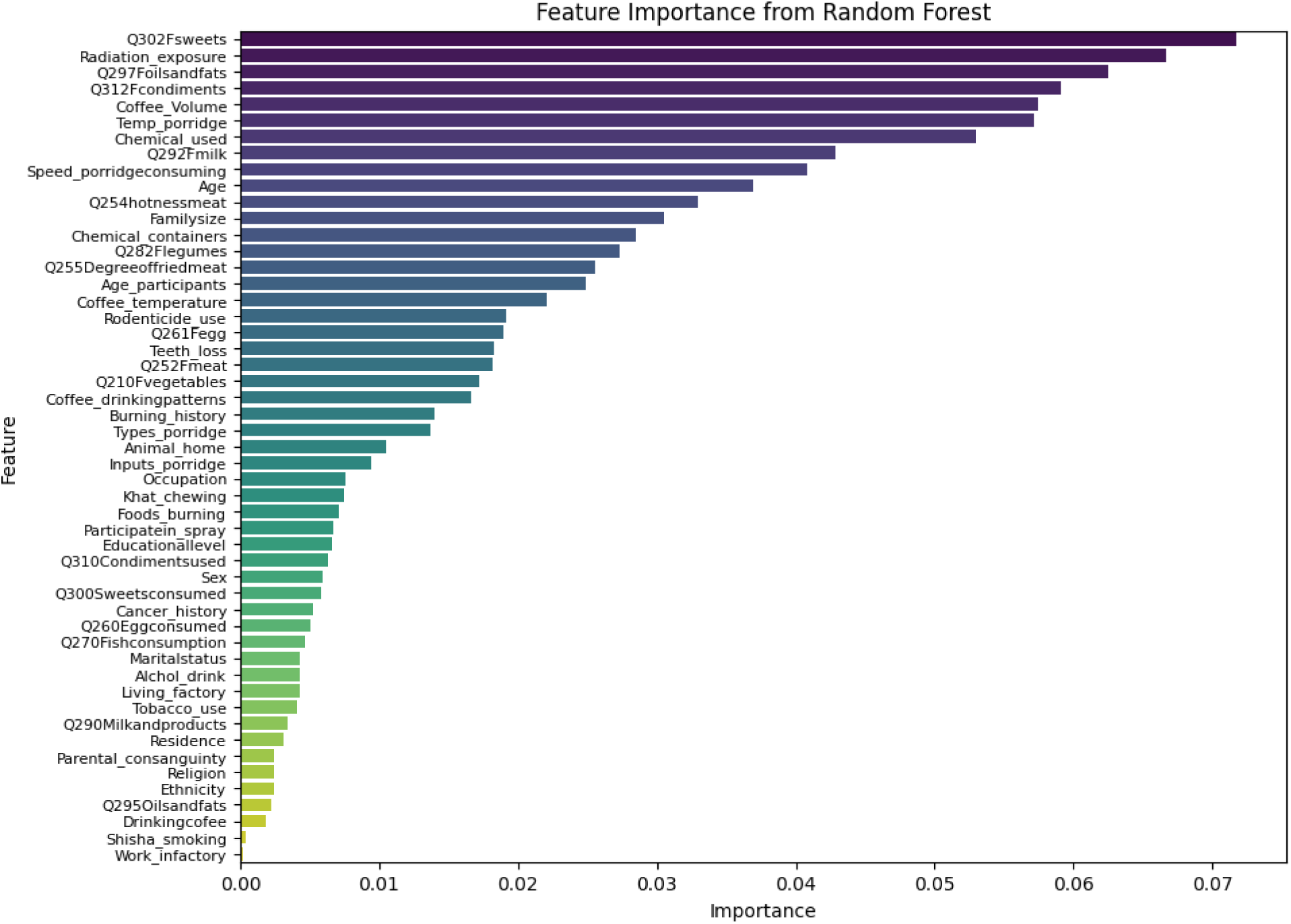
The feature importance profile from the ensemble feature ranking model highlights a cluster of dietary and environmental variables as the most influential predictors.

Several studies [44, 45] have reported positive associations between preserved or high-salt foods, sugary items, and fried or fatty foods and increased risk of esophageal squamous cell carcinoma (ESCC). Likewise, the prominence of coffee volume, temperature of porridge, and variables reflecting chemical use mirrors robust evidence that consumption of very hot foods or beverages significantly increases esophageal cancer risk, with multiple meta-analyses demonstrating two- to three-fold higher odds among individuals regularly consuming foods or drinks at high temperatures. These parallels suggest that the model is capturing biologically plausible and literature-supported associations, particularly those related to thermal injury, dietary irritants, and environmental exposures [3].

Features appearing lower in the importance spectrum, although contributing less individually, still reflect meaningful behavioral and demographic contexts observed in many esophageal cancer studies. Variables such as “burning history,” “types of porridge,” “animal home,” “occupation,” and “khat chewing” signal lifestyle and environmental conditions that may shape underlying exposure profiles, even if their direct predictive power is modest in the model.

Socio-demographic indicators, including “education level,” “sex,” “marital status,” “ethnicity,” and “residence,” also play secondary roles, which is consistent with literature showing that while these factors may modify risk or influence exposure pathways, they seldom exert effects as strong as dietary patterns or environmental hazards [46]. Taken together, the feature-importance distribution underscores a multifactorial structure in which dietary behaviors, food-handling practices, and environmental exposures form the core predictive signal, supported by a broader constellation of lifestyle and demographic variables that provide contextual nuance rather than driving the model’s predictions directly.

In this study, we conducted two experiments to generate reduced feature sets by progressively removing predictors with the lowest feature-importance scores, as illustrated at the bottom of Fig. 3.

In Experiment 1, the last eight features, “Residence,” “Parental consanguinity,” “Ethnicity,” “Religion,” “Q295Oilsandfats,” “Drinkingcoffee,” “Shisha_smoking,” and “Work_infactory”, were removed. In Experiment 2, only the last two features, “Shisha_smoking” and “Work_infactory”, were excluded.

This strategy is motivated by the clear pattern observed in the feature-importance ranking, where several socio-demographic variables such as education level, sex, marital status, ethnicity, and place of residence consistently fall within the lower half of the importance spectrum. Such variables may provide contextual background but do not substantially influence the model’s predictive behavior compared to the more dominant dietary, behavioral, and environmental risk factors.

By systematically excluding these low-impact features, the two experiments allow us to evaluate whether model performance can be preserved or even improved while reducing dimensionality and enhancing interpretability. In the subsequent analysis, we select the two highest-performing experimental settings and present the remaining results using a comparative framework, enabling a clear assessment of how differences in feature-reduction strategies affect classification performance across models.

After selecting the most important features, several ensemble models were developed based on the proposed ensemble classification framework (Fig. 2). The performance of the best proposed esophageal cancer detection model was evaluated using an averaged confusion matrix across all model components in Experiment 2 (Fig. 5).

The model correctly identified 41 esophageal cancer cases (true positives) and 21 non-cancer cases (true negatives). A single false-positive prediction was observed, while no false-negative cases were detected. Of particular clinical importance of Beta Risk (Type II error probability) and Diagnostic Sensitivity, the ensemble HGBC model exhibited zero false negatives, corresponding to a beta risk (β) of 0. Accordingly, the statistical power (1 − β) of the model was 100%, indicating that all true esophageal cancer cases present in the evaluation dataset were successfully detected. This outcome reflects perfect sensitivity within the tested cohort.

In the context of cancer diagnostics, beta risk represents the probability of failing to detect disease when it is present, a scenario that carries significant clinical consequences due to delayed diagnosis and treatment. The absence of false-negative predictions in this evaluation suggests that the proposed model effectively mitigates this risk, aligning with the clinical priority of maximizing sensitivity in oncological decision-support systems. While one false-positive prediction was observed, this trade-off is generally considered acceptable in decision-support systems, where minimizing missed cancer cases outweighs the cost of additional diagnostic follow-up.

The ROC curve for the ensemble HGBC model using a full (left panel) and reduced feature set (right panel) in Experiment 2 is shown in Fig. 4 which shows both having an AUC value of 0.994. The ROC curve for the ensemble HGBC model using a reduced feature set in Experiment 2 shown in Fig. 6 demonstrates a pronounced ability to discriminate between the two classes, as reflected by its steep initial ascent and its near-horizontal trajectory across the upper region of the plot. This shape indicates that the model achieves high true positive rates while maintaining low false positive rates, a desirable characteristic in classification tasks where minimizing missed detections and limiting false alarms are both critical. The curve’s proximity to the top-left corner of the ROC space further underscores the model’s effectiveness, suggesting that even with a simplified feature representation, the classifier captures meaningful patterns that separate the classes with high fidelity.

**Figure 4.**
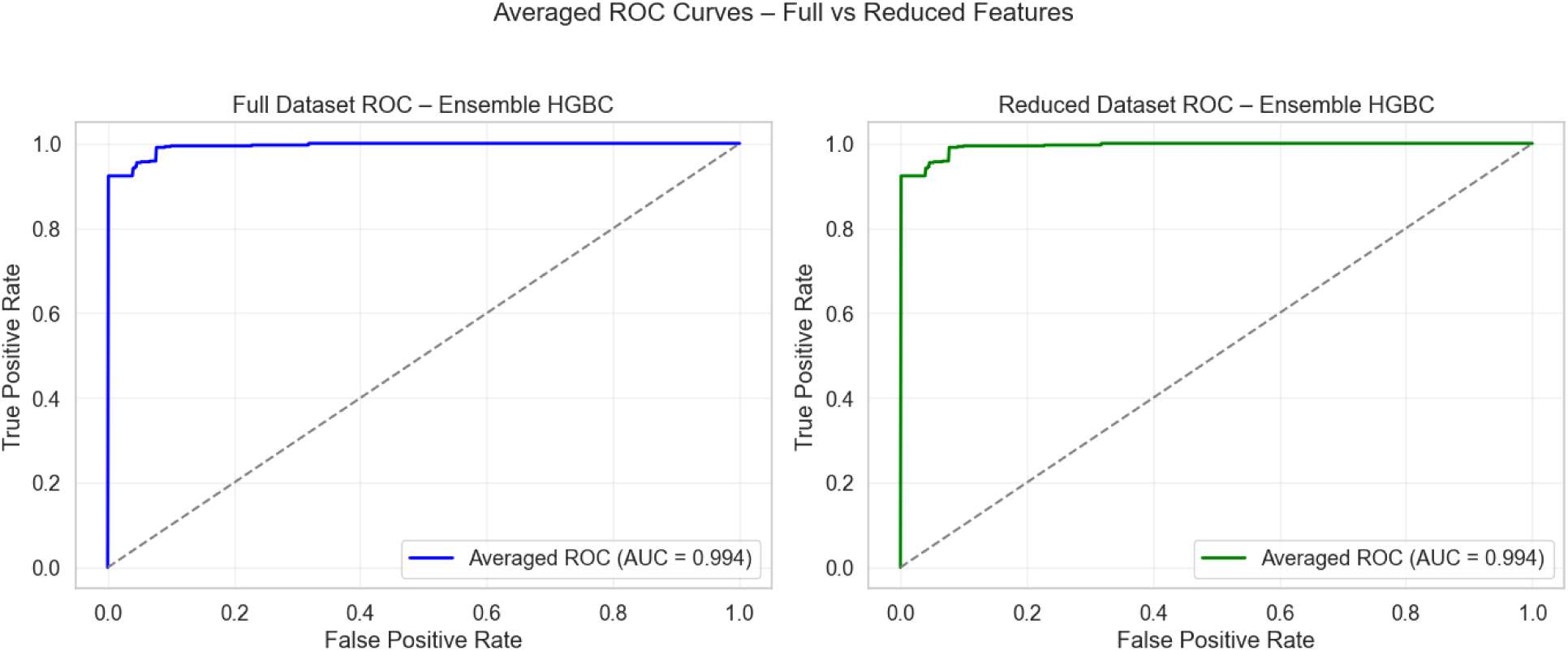
ROC for the ensemble HGBC model with full and reduced features in the data set in Experiment 2.

**Figure 5.**
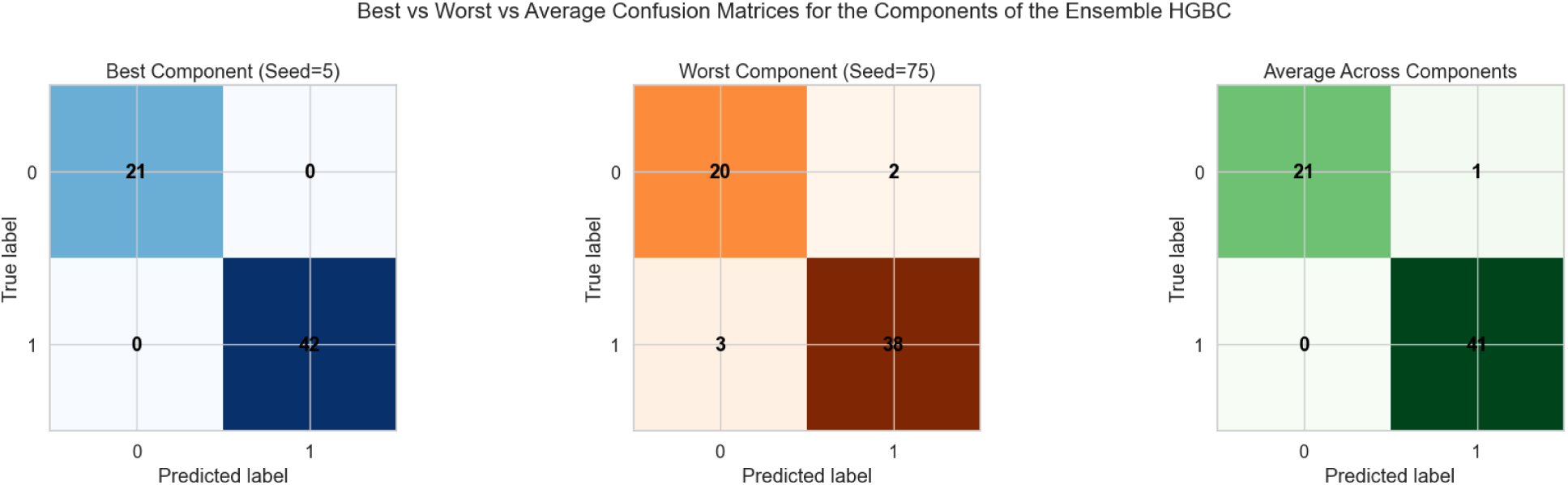
Confusion matrix of the ensemble HGBC model with reduced features in the data set in Experiment 2.

**Figure 6.**
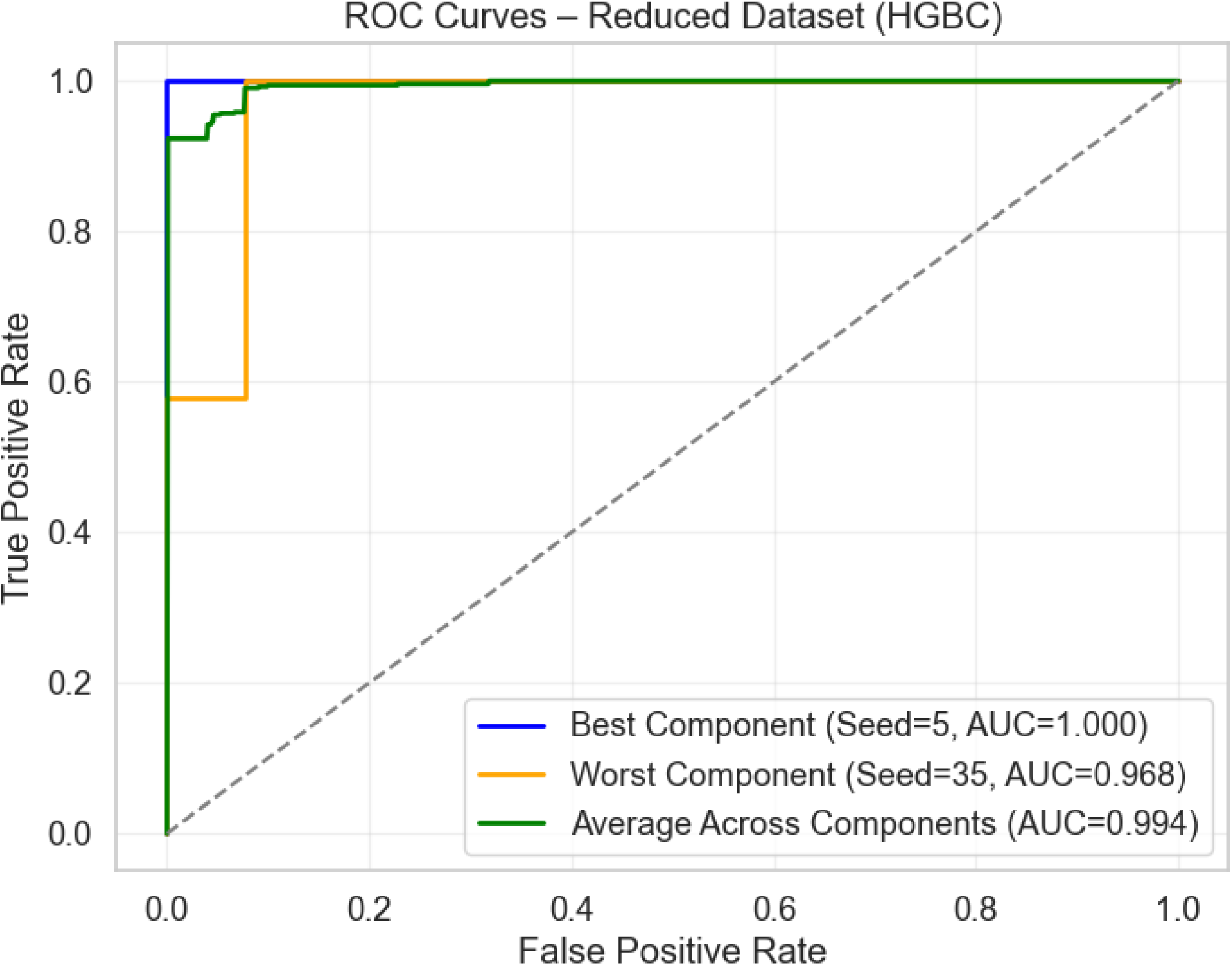
ROC for the ensemble HGBC model with reduced features in the data set in Experiment 2.

An AUC of 0.994, as illustrated in Fig. 6, highlights the ensemble HGBC model’s exceptional discriminative ability, reinforcing the visual impression of strong performance and positioning it well within the range typically indicative of an excellent classifier. Such a high AUC indicates that the reduced-feature version of the model retains nearly all of the predictive power expected from more complex configurations, implying that key information captured by the hemoglobin-related variable(s) may be highly informative for the underlying classification task. This finding not only highlights the robustness and efficiency of the model but also suggests practical advantages, such as reduced computational overhead, improved interpretability, and potential clinical or operational feasibility without substantial loss of accuracy.

The performance of the five ensemble design based on the machine-learning methods we have briefly discussed in Sec. 3.3, namely, Voting-Based Histogram Gradient Boosting Ensemble (Model I), Voting-Based Extreme Gradient Boosting Ensemble (Model II), Voting-Based Extreme AdaBoost (Model III), Voting-Based Random Forest Ensemble (Model IV), and Voting-Based K-Nearest Neighbors Ensemble (Model V), was evaluated using the processed dataset consisting of 312 individuals, of whom 104 were confirmed esophageal cancer (EC) cases and 208 were non-cancer controls. Each instance contained 52 selected features, which we refer to as “full,” representing demographic characteristics, clinical symptoms, tumor-related factors, and other medically relevant variables derived through feature-importance filtering (Section 3). The binary classification task therefore involved distinguishing EC = 1 from non-EC = 0, and model outputs were assessed relative to these labels.

A comparison of the two experiments presented in Tables 1 and 2 based on ensemble-based machine learning models using reduced and full feature sets demonstrates that the strongest classifiers, namely Model I, Model II, and Model III maintain consistently high performance across both experimental setups, with only minor variations attributable to differences in feature selection. Overall, the results indicate that feature reduction does not degrade predictive performance and, in several cases, leads to marginal improvements.

**Table 1.**
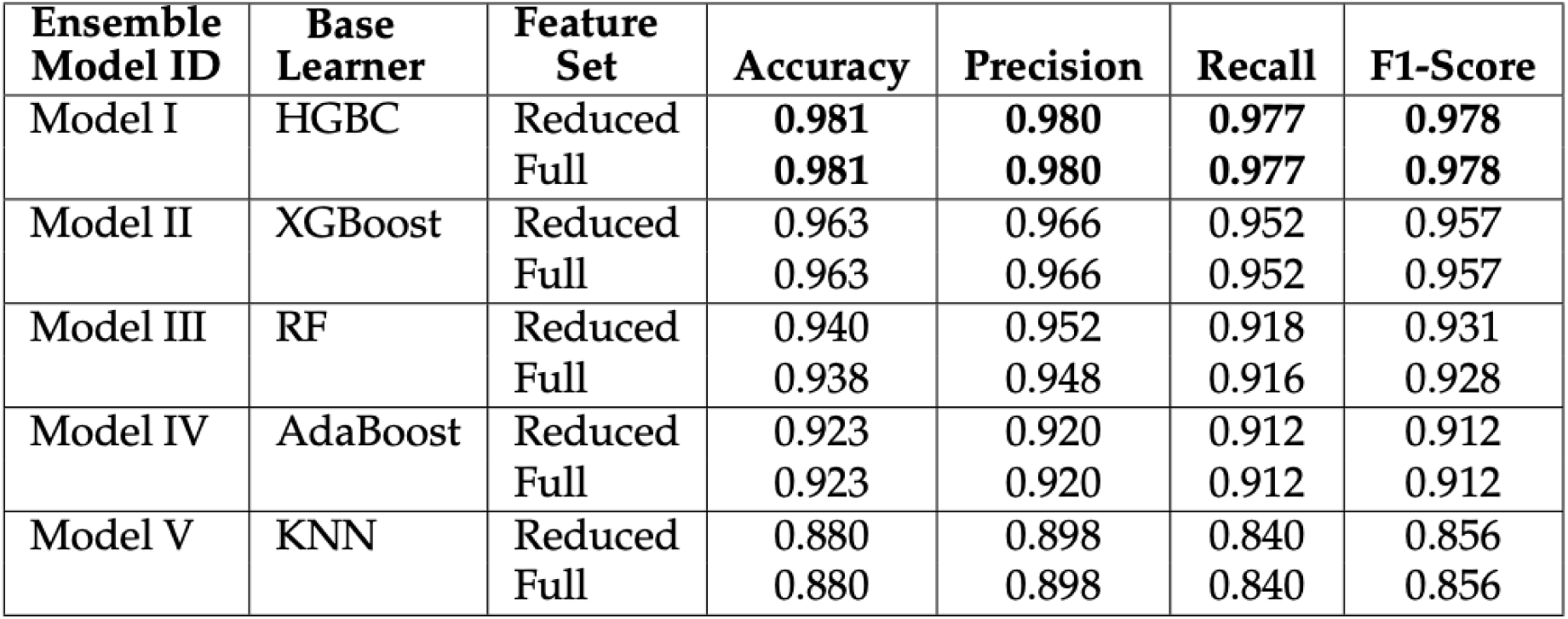
Performance comparison of ensemble-based machine learning models using reduced and full feature sets in Experiment 1.

| **Ensemble Model ID** | **Base Learner** | **Feature Set** | **Accuracy** | **Precision** | **Recall** | **F1-Score** |
| --- | --- | --- | --- | --- | --- | --- |
| Model I | HGBC | Reduced | **0.981** | **0.980** | **0.977** | **0.978** |
|  |  | Full | **0.981** | **0.980** | **0.977** | **0.978** |
| Model II | XGBoost | Reduced | 0.963 | 0.966 | 0.952 | 0.957 |
|  |  | Full | 0.963 | 0.966 | 0.952 | 0.957 |
| Model III | RF | Reduced | 0.940 | 0.952 | 0.918 | 0.931 |
|  |  | Full | 0.938 | 0.948 | 0.916 | 0.928 |
| Model IV | AdaBoost | Reduced | 0.923 | 0.920 | 0.912 | 0.912 |
|  |  | Full | 0.923 | 0.920 | 0.912 | 0.912 |
| Model V | KNN | Reduced | 0.880 | 0.898 | 0.840 | 0.856 |
|  |  | Full | 0.880 | 0.898 | 0.840 | 0.856 |

**Table 2.** Performance comparison of ensemble-based machine learning models using reduced and full feature sets in Experiment 2.

| Ensemble Model ID | Base Learner | Feature Set | Accuracy | Precision | Recall | F1-Score |
| --- | --- | --- | --- | --- | --- | --- |
| Model I | HGBC | Reduced | **0.983** | **0.982** | **0.980** | **0.981** |
|  |  | Full | 0.980 | 0.980 | 0.977 | 0.978 |
| Model II | XGBoost | Reduced | 0.961 | 0.966 | 0.946 | 0.955 |
|  |  | Full | 0.963 | 0.966 | 0.952 | 0.957 |
| Model III | RF | Reduced | 0.940 | 0.951 | 0.917 | 0.930 |
|  |  | Full | 0.938 | 0.948 | 0.916 | 0.928 |
| Model IV | AdaBoost | Reduced | 0.924 | 0.921 | 0.913 | 0.914 |
|  |  | Full | 0.923 | 0.920 | 0.912 | 0.912 |
| Model V | KNN | Reduced | 0.896 | 0.914 | 0.859 | 0.877 |
|  |  | Full | 0.880 | 0.898 | 0.840 | 0.856 |

Model I consistently emerges as the top-performing algorithm across both experiments. In Experiment 1, Model I achieves identical peak accuracies of 0.981 for both the full and reduced feature sets, while in Experiment 2 the reduced-feature configuration attains the highest overall accuracy of 0.983. Similar trends are observed for precision, recall, and F1-score, confirming Model I’s robustness to feature elimination and its strong ability to capture complex nonlinear relationships. These results highlight the suitability of Model I for high-stakes predictive tasks, where stability and reliability are critical.

Model II also demonstrates stable and competitive performance across both experiments. In Experiment 1, Model II achieves identical accuracies of 0.963 for the full and reduced feature sets, indicating insensitivity to feature reduction. In Experiment 2, a slight reduction in accuracy is observed for the reduced feature set (0.961) compared to the full set (0.963), though the difference is negligible. The consistency of Model II across experiments reflects its inherent regularization mechanisms and resilience to redundant or correlated features.

Model III exhibits slightly greater sensitivity to feature selection compared with Model I and Model II. In both experiments, the reduced-feature Model II achieves an accuracy of 0.940, marginally outperforming the full-feature model (0.938). This suggests that removing less informative variables improves Model II stability by reducing noise and overfitting while preserving strong predictive capability.

Among the mid-tier classifiers, Model IV and Model V display more noticeable effects of feature reduction. In Experiment 1, both models exhibit identical performance across full and reduced feature sets, with accuracies of 0.923 for Model IV and 0.880 for Model V. In Experiment 2, however, the reduced-feature versions outperform their full-feature counterparts, with Model V improving from 0.880 to 0.896 accuracy and Model IV showing a modest gain from 0.923 to 0.924. These findings suggest that distance-based and boosting-based methods benefit from the removal of irrelevant or noisy features, which can otherwise distort neighborhood relationships or weak learner updates.

Overall, the comparative analysis across Experiments 1 and 2 confirms that feature reduction preserves predictive performance across all evaluated models, with negligible differences in accuracy, precision, recall, and F1-score. In several cases, reduced feature sets slightly enhance performance, indicating that the selected features retain the most informative variables while eliminating redundancy.

Across both experiments, ensemble learning approaches consistently outperform or match simpler classifiers. Ensemble of the boosting-based ML models, particularly Model I and Model II, deliver the highest and most balanced performance, demonstrating strong generalization ability even under reduced feature representations. These results confirm that the proposed ensemble models are well suited for esophageal cancer prediction in this dataset, providing high accuracy alongside clinically relevant sensitivity and robustness.

Typically, ensemble machine-learning models trained on a reduced dataset show lower predictive accuracy because removing instances or features leads to information loss and weaker data representation [47, 48]. However, in our case, the models trained on the feature-reduced subset achieved performance measures that were statistically comparable to, or even better than, those trained on the full dataset.

This outcome provides strong empirical support for the proposed ensemble random forest feature-ranking framework. The framework effectively identifies the most relevant and non-redundant features, indicating that the removed features contribute little or even introduce noise to the prediction process. As a result, the smaller feature subset still preserves the essential discriminative information required for constructing a high-performing predictive model.

Usually ensemble models enhance machine learning performance by aggregating multiple base learners into a final decision that leverages complementary model outputs to improve predictive reliability. Our proposed ensemble approach reduces sensitivity to minor perturbations in training data and hyperparameter configurations, thereby increasing stability and generalization capacity. As a result, ensembles exhibit diminished vulnerability to noise and inherent variability within datasets and model configuration. Although improvements in accuracy may sometimes be incremental, and in certain contexts even limited, the associated reduction in variance and enhancement of consistency are critical, particularly in high-stakes domains such as medical diagnostics, where dependable and reproducible predictions are essential for safe clinical decision-making.

The comparison plots in Fig. 7 indicate that the proposed ensemble models consistently achieve high and stable accuracy across both full and reduced feature sets, with boosting-based methods showing the strongest overall performance. Ensemble model designed based on HGBC algorithm attains accuracies of approximately 0.98 in both experiments, including the reduced feature configuration, while ensemble model designed based on XGBoost algorithm remains similarly stable at around 0.96 across feature sets and experimental setups. Ensemble model designed based on RF achieves accuracies close to 0.94 in both cases, with a slight and consistent advantage for the reduced feature set. Ensemble model designed based on KNN records an accuracy of about 0.88 in Experiment 1 for both feature sets, improving to approximately 0.90 with reduced features in Experiment 2, indicating sensitivity to dimensionality. Ensemble model designed based on AdaBoost remains stable at roughly 0.92 across both experiments, with minimal differences between full and reduced features. Overall, these results demonstrate the robustness and effectiveness of ensemble models, particularly boosting-based approaches, and show that reduced feature sets can maintain comparable accuracy while improving computational efficiency.

**Figure 7.**
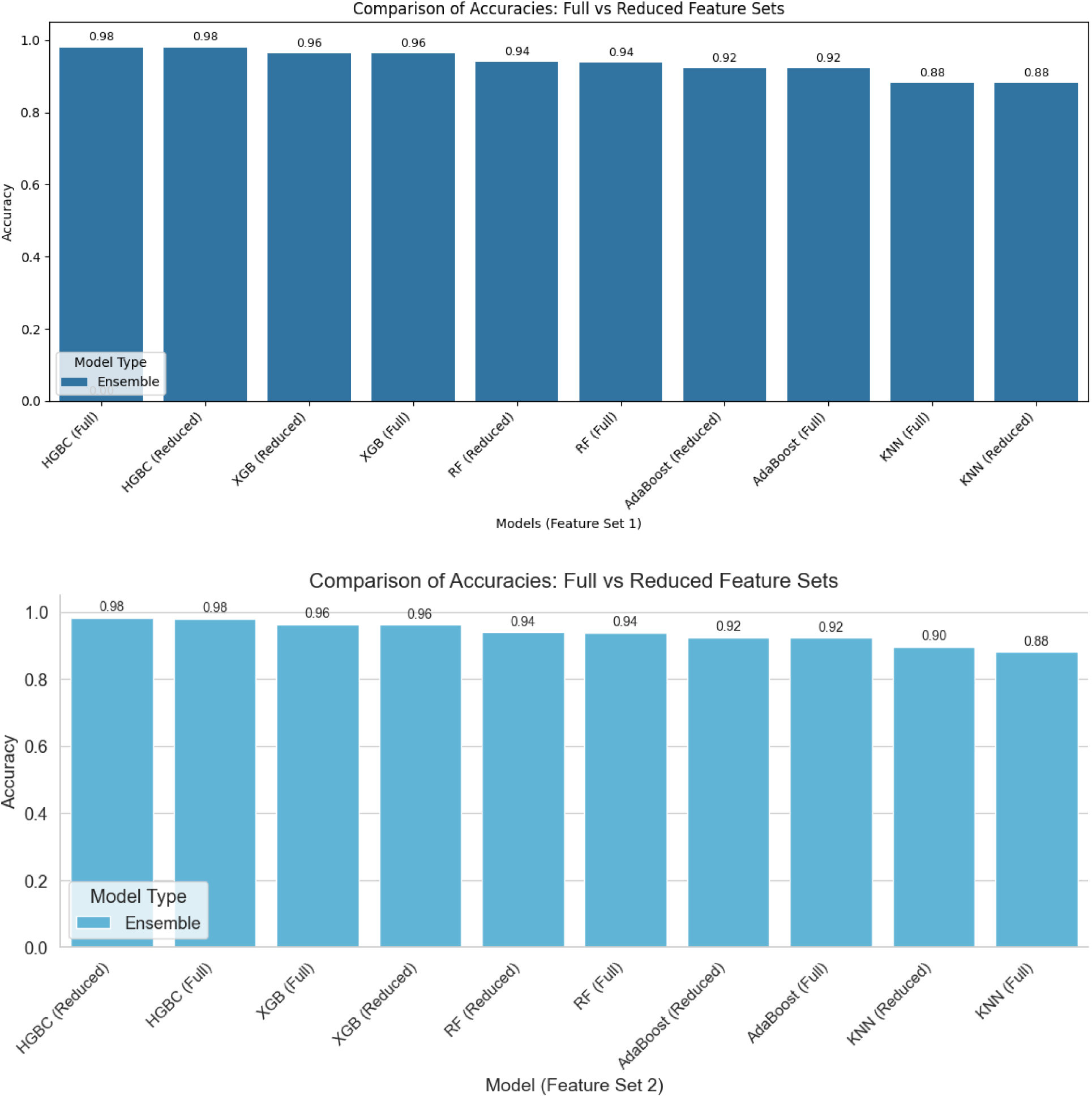
Accuracy comparison for the evaluated ensemble-based machine learning models between Experiment 1 (top panel) and Experiment 2 (bottom panel).

## 6. Conclusion

This study demonstrates the effectiveness of ensemble-based machine learning approaches, particularly ensemble of boosting-based models, for improving the detection and risk stratification of esophageal cancer (EC) using structured clinical, behavioral, dietary, and environmental data. The proposed framework integrates a multi-seed ensemble classification strategy with an ensemble-based feature-ranking approach, evaluated on both full and reduced feature sets. The findings show that a reduced subset of features, primarily related to dietary practices, consumption of hot foods, environmental exposures, and selected behavioral factors, is sufficient to preserve, and in some cases enhance, predictive performance across multiple classifiers. This underscores the critical role of these factors in EC risk assessment.

Among the evaluated models, Model I consistently achieved the highest and most stable performance across both experiments. In Experiment 1, Model I attained an accuracy of 0.981 for both full and reduced feature sets, while in Experiment 2 the reduced-feature model achieved the highest overall accuracy of 0.983, accompanied by strong precision, recall, and F1-score values. These results highlight Model I’s robustness to feature reduction and its ability to effectively model complex nonlinear relationships inherent in EC risk data.

Model II also demonstrated consistently strong performance, maintaining an accuracy of 0.963 across full and reduced feature sets in Experiment 1, with only a marginal decrease to 0.961 for the reduced-feature model in Experiment 2. The stability of Model II under feature reduction reflects its inherent regularization and feature-handling capabilities.

Model III showed slightly greater sensitivity to feature selection; however, the reduced-feature configuration marginally outperformed the full-feature model in both experiments, indicating that dimensionality reduction can improve model stability by minimizing noise and redundancy.

Overall, the results demonstrate that Model I achieves zero beta risk, maximal statistical power, and high diagnostic sensitivity for esophageal cancer detection in the evaluated dataset. These findings support its potential suitability for assistive diagnostic use. Further validation on larger, independent, and multi-center datasets is necessary to assess generalizability and confirm performance stability.

While these results are promising, several limitations remain. The dataset represents a specific high-risk geographic region and has a moderate sample size, which may limit external generalizability. Future work should expand the dataset, integrate additional modalities such as imaging or genomic features, and explore explainable AI techniques to enhance clinical interpretability. Incorporating deep learning models or hybrid architectures may further improve predictive capabilities.

In conclusion, this study presents a robust and interpretable framework for esophageal cancer classification and demonstrates the practical value of ensemble machine learning models in supporting early detection and clinical decision-making. The findings highlight the potential of reduced-feature ensemble models to deliver accurate, efficient, and clinically relevant predictions, particularly in resource-constrained healthcare settings, thereby contributing to improved patient outcomes and advancing precision medicine for esophageal cancer.

## Author Contributions

Conceptualization, M.G., S.C., and R.M.; methodology, S.C.; software, S.C. and R.M.; validation, M.G., R.M., S.C., and H.D.; formal analysis, M.G., R.M., and S.C.; investigation, M.G., R.M., and S.C.; data curation, H.D. and A.A.; writing, original draft preparation, M.G.; writing, reviewing and editing, R.M., G. A. and A.A.; visualization, M.G.; supervision, S.C.; project administration, R.M. and S.C. All authors have read and agreed to the published version of the manuscript.

## Informed Consent Statement

Informed consent was obtained from all subjects involved in the study.

## Data Availability Statement

The dataset is owned by H.D. and may be made available from the corresponding author upon reasonable request.

